# Understanding the potential contribution of polygenic risk scores to the prediction of gestational and type 2 diabetes in women from British Pakistani and Bangladeshi groups: a cohort study in Genes and Health

**DOI:** 10.1101/2025.01.15.25320604

**Authors:** Julia Zöllner, Binur Orazumbekova, Sam Hodgson, Genes and Health Research Team, David A. van Heel, Stamatina Iliodromiti, Moneeza Siddiqui, Rohini Mathur, Sarah Finer, Jen Jardine

## Abstract

**Background:** British Pakistani and Bangladeshi women have disproportionately high rates of gestational diabetes mellitus (GDM), with prevalence estimates up to three times higher than in the general population. They are also at increased risk of progressing to type 2 diabetes, leading to significant health complications. Despite this, predictive models tailored to this high-risk, yet understudied group are lacking.

**Objectives:** To investigate whether combining genetic and traditional clinical data improves risk prediction of GDM and progression to type 2 diabetes among British Pakistani and Bangladeshi women. We hypothesized that incorporating polygenic risk scores (PRS) would enhance the predictive accuracy of existing models.

**Study Design:** An observational cohort study utilizing the Genes & Health dataset, which includes comprehensive electronic health records. Women who gave birth between 2000 and 2023, both with and without a history of GDM, were included. Controls were defined as women without a GDM diagnosis during this period but who had a birth record. A total of 117 type 2 diabetes or GDM PRS were tested to determine the optimal PRS based on predictive performance metrics. The best-performing PRS was integrated with clinical variables for statistical analyses, including descriptive statistics, chi-square tests, logistic regression, and receiver operating characteristic curve analysis.

**Results:** Of 13,489 women with birth records, 10,931 were included in the analysis, with 29.3% developing GDM. Women with GDM were older (mean age 31.7 years, p < 0.001) and had a higher BMI (mean 28.4 kg/m², p < 0.001) compared to controls. The optimal PRS demonstrated a strong association with GDM risk; women in the highest PRS decile had significantly increased odds of developing GDM (OR 5.66, 95% CI [4.59, 7.01], p = 3.62 × 10⁻). Furthermore, the risk of converting from GDM to type 2 diabetes was 30% in the highest PRS decile, compared to 19% among all GDM cases and 11% in the lowest decile. Incorporating genetic risk factors with clinical data improved the C-statistic for predicting type 2 diabetes following GDM from 0.62 to 0.67 (pL=L4.58L×L10⁻L), indicating better model discrimination.

**Conclusion:** The integration of genetic assessment with traditional clinical factors significantly enhances risk prediction for British Pakistani and Bangladeshi women at high risk of developing type 2 diabetes after GDM. These findings support the implementation of targeted interventions and personalized monitoring strategies in this high-risk population. Future research should focus on validating these predictive models in external cohorts and exploring their integration into clinical practice to improve health outcomes.

**AJOG at a glance:**

- *Why was this study conducted?* To understand the contribution of polygenic risk scores in predicting gestational diabetes and subsequent type 2 diabetes in women of British Pakistani and Bangladeshi descent.
- *What are the key findings?* Women with higher genetic risk scores were found to have a significantly increased likelihood of developing gestational diabetes and type 2 diabetes.
- *What does this study add to what is already known* This research enhances understanding of how genetic factors can improve risk prediction in high-risk populations, potentially informing targeted preventive strategies.

## Introduction

Gestational diabetes, or diabetes which newly arises in pregnancy, is a substantial risk factor for type 2 diabetes, with women who have gestational diabetes having at least a seven-fold increase of type 2 diabetes compared to women who have a pregnancy without gestational diabetes.[1,2] Understanding the relationship between gestational and type 2 diabetes not only provides important insights into each disease, but also opportunities to develop novel interventions to reduce the risk of progression to type 2 diabetes.

This risk of progression is particularly high in women of South Asian ethnic origin, of whom approximately 35% will have impaired glucose tolerance at 6-8 weeks postpartum.[3] People of South Asian ethnic origin are at elevated risk overall of type 2 diabetes, which occurs younger, at a lower BMI and with fewer comorbidities than in Caucasian populations. [4] Targeting this population is therefore a high priority to reduce the overall incidence of type 2 diabetes.

Recent studies have demonstrated that incorporating genetic as well as demographic and disease information can improve the accuracy of prediction of likelihood of type 2 diabetes in women with GDM, including those of South Asian ethnic origin.[5] Many of the genetic and environmental risk factors associated with GDM are shared with type 2 diabetes.[6] Women diagnosed with GDM often have a higher likelihood of having at least one parent with type 2 diabetes compared to those with normal gestational glycaemia[7] and polygenic risk scores (PRS) for type 2 diabetes demonstrate correlation with the risk of developing GDM.[8] Pregnancy offers an opportunity to combine PRS with non-genetic risk factors, allow an early risk assessment to offer preventive interventions and/or medicines. To date only a few small studies have investigated polygenic risk in GDM demonstrating linear correlation between those in the highest risk deciles and glucose intolerance.[9,10] No studies have integrated a polygenic risk combined with pregnancy risk factors in a large real-world dataset like Genes and Health to improve clinical prediction. In this study, we aim to understand the demographic, disease and genetic associations for gestational diabetes and subsequent development of type 2 diabetes among women of South Asian ethnic origin.

## Materials and Methods

This is an observational cohort study using the Genes & Health (G&H) cohort of people of British Pakistani and Bangladeshi (BPB) ethnic origin. The population of interest are women who have had a recorded registerable birth between 2000 and 2023 with a history of gestational diabetes.

### Data source and study population

Genes and Health recruits BPB people aged 16 years and above, predominantly from community and primary care settings. Further detail about the cohort has been previously described.[6] We used the 2023 data release (secondary care data May 2023, primary care data November 2023), which comprised electronic health record (EHR) data from primary and secondary care, and genotype data from the Illumina Infinium Global Screening Array V3 Chip[11] Descriptions of quality control, imputation of genotype data, as well as filtering and principal component analysis are provided in Supplementary Methods.

Women from the G&H population were included if they had a SNOMED or ICD-10 code (Supplementary Table 1) indicating that they had given birth on or after 1^st^ April 2000 and on or before the latest EHR data refresh.[12] Births prior to this year were excluded a priori as the quality of birth records in electronic health data in England improved from 2000.

#### Determination of exposures and outcomes

For this study, the primary exposure was gestational diabetes, and the primary outcome was development of type 2 diabetes following pregnancy. We identified women as having gestational diabetes if they had a code for gestational diabetes (Supplementary Table 1) within the six months preceding or six weeks following a birth episode in either HES or the GP record from the year 2000 onwards. We additionally inspected women with a code of type 2 diabetes in this period. If diabetes was only coded in pregnancy and the postpartum period up to 3 months, and only treated with metformin and insulin (as recommended by the National Institute for Clinical Excellence[13]) then this was considered to indicate a diagnosis of gestational diabetes. If diabetes was coded or treatment prescribed more than six months before birth, or after 3 months postpartum, and/or if suphonylureas were prescribed, then the record was treated as indicating diabetes outside of pregnancy.

The following additional filtering approaches were applied for mothers with multiple pregnancy records in order to keep a single pregnancy per mother for analysis: (i) if GDM was not diagnosed at any of the pregnancies, phenotype data at the latest available time point was kept (i.e., keep older women without GDM), (ii) if GDM was diagnosed during any of the pregnancies included in the study, the earliest time point where GDM was diagnosed was kept (i.e., keep younger women with GDM). When assessing parity – we used GDM diagnosis dates and birth record dates to assess which came first. Due to higher rates of relatedness in this specific population, related individuals by third degree or closer were removed from the analysis.

Covariates included age at time of giving birth, BMI closest to or during pregnancy, parity (nulliparous or multiparous), and ethnicity (Bangladeshi or Pakistani). These were used to derive an epidemiological model.

#### Choice of polygenic risk score

At the time of the analysis there were no South Asian-specific type 2 diabetes PRS data available, therefore type 2 diabetes (n=115) and GDM (n=2) polygenic risk scores available on the PGS catalogue were compared against performance in our dataset (Supplementary Figure 1). In addition, weights were derived from the DIAGRAM’s 2022 multi-ethnic type 2 diabetes GWAS meta-analysis, which included 8.3% South Asians (51.1% European, 26.4% East Asian, 6.6% African ancestry and 5.6% other ethnic backgrounds).[14] Finally, weights were derived from the largest and most recent GDM GWAS (100% European – Finland and Estonia).[15] PGS002771 (100% European) performed the best and was used for subsequent analysis; the derivation of this PRS is described elsewhere.[16]

#### Statistical analysis

Participant characteristics were evaluated using descriptive statistical methods. For continuous variables that followed a normal distribution, the mean and standard deviation (SD) are presented. For variables that were not normally distributed, the median and interquartile range (IQR) are also included. Categorical variables are summarized as proportions. A two-sided p-value of less than 0.05 was considered statistically significant. Data analysis was performed on the Google Cloud Trusted Research environment. For statistical analysis RStudio version 2023.09.1 was used.

#### Missing data

Missing data was imputed using the R program Mice. We imputed BMI values for 1010 volunteers (763 controls, 247 cases) and parity for 394 cases using multiple imputation with m=10. Estimates were pooled using Rubin’s rules[17].

#### Ethical approval

ELGH (East London Genes and Health) operates under ethical approval, 14/LO/1240, from London South East NRES Committee of the Health Research Authority, dated 16 September 2014. This protocol was approved by ELGH on 5^th^ July 2023, reference S00099.

## Results

The Genes and Health dataset includes 59,150 participants, of whom 32,494 are recorded as women. Of these women, 13,489 had a record of giving birth after the 1^st^ January 2000 in either their primary or secondary care record; 27,681 discrete birth episodes were identified. Women who had a birth record were generally younger (controls median year of birth 1982 [IQR 1977-1987]; cases mean 1983 [IQR 1979-1987]) compared to participants who had no record of having given birth (median 1979[IQR 1967-1990]. Median number of births in the cohort was two per woman.

10931 women were included in the analysis (Figure 1). The demographics of the included cases and controls are described in Table 1. 29.3% of women in the cohort developed GDM in their included pregnancy. Women who developed GDM were slightly older (mean age 31.7 compared to 31.3 years, p<0.001) and had a higher BMI (mean BMI 28.4 compared to 27.3kg/m^2^, p<0.001), and more likely to be Bangladeshi than women who did not have GDM (75.3% of all women with GDM were Bangladeshi compared to 59.7% of all women without GDM, p<0.001). Nulliparous women were more likely to be represented in the GDM group (52.7% compared to 29.5%, p<0.01).

**Figure 1.**
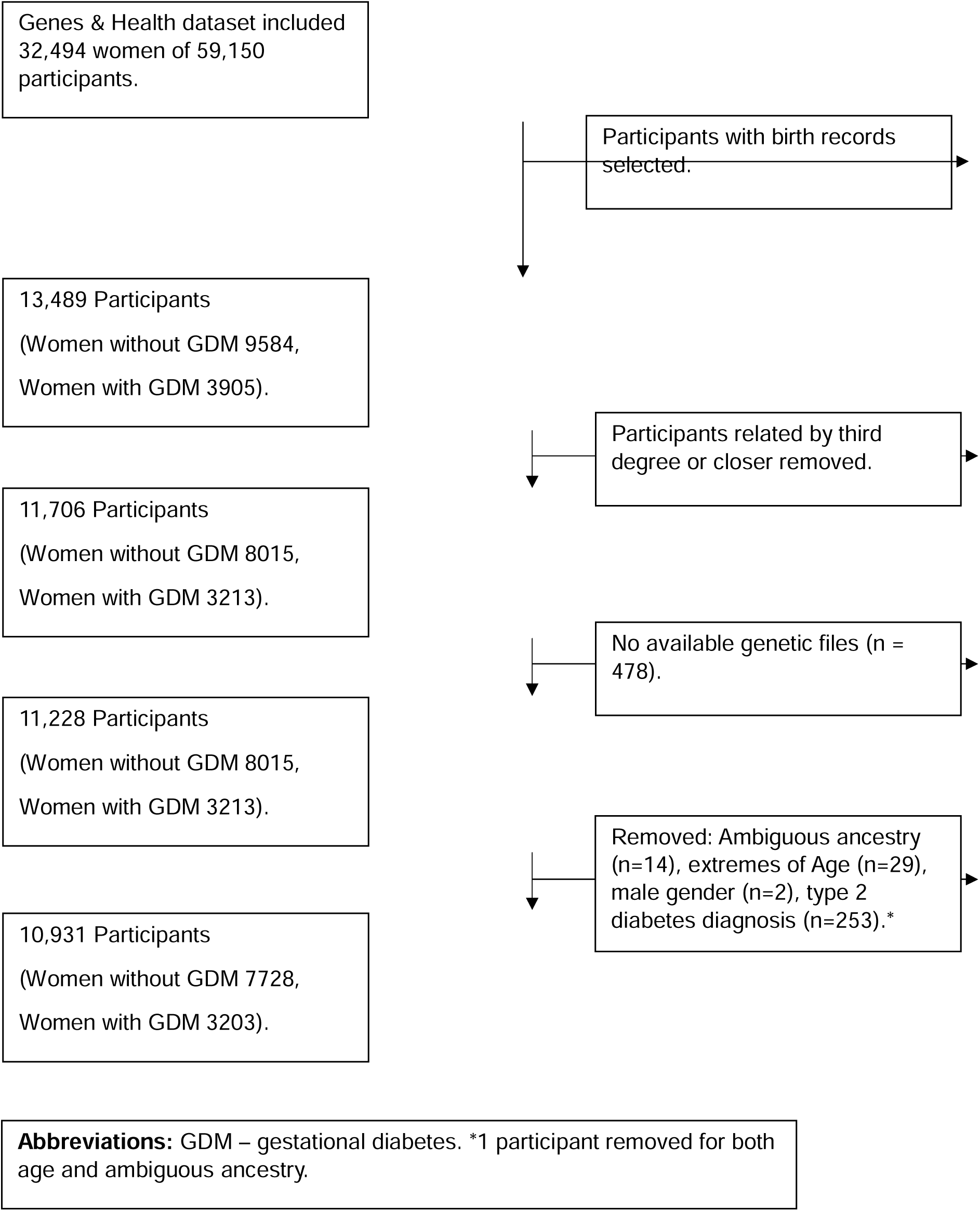
Strobe chart.

**Table 1.** Characteristics of 10,931 Genes and Health participants who gave birth after 1^st^ April 2000.

| Characteristic | Women who did not have GDM (n, %) | Women who had GDM (n, %) | p value |
| --- | --- | --- | --- |
| All women | 7728 (70.70%) | 3203 (29.30%) | . |
| Age (years), mean (SD) | 31.36 (4.92) | 31.73 (5.24) | <0.001 |
| BMI, mean (SD)* | 27.33 (5.14) | 28.47 (5.19) | <0.001 |
| Parity at included birth, n (%)** |  |  |  |
| Nulliparous | 2059 (29.58) | 1373 (52.75) | <0.001 |
| Multiparous (1 or more) | 4902 (70.42) | 1230 (47.25) |  |
| Genetic ancestry, n (%) |  |  |  |
| British Bangladeshi | 4611 (59.7) | 2412 (75.3) | <0.001 |
| British Pakistani | 3117 (40.3) | 791 (24.7) |  |
| Polygenic risk score for T2DM, n (%) |  |  |  |
| 1<sup>st</sup> Decile | 949 (12.28) | 145 (4.53) | <0.001 |
| 10<sup>th</sup> Decile | 586 (7.58) | 507 (15.83) |  |

Characteristics of participants (cases and non-cases) included in the analysis. Data is presented as means (standard deviation) unless otherwise stated. P-values are calculated from Chi-squared test for categorical variables and independent t-test for continuous variables. GDM diagnosis is made based on coding in primary and secondary care records. Abbreviations: GDM – gestational diabetes; SD – standard deviation; T2DM – type 2 diabetes. \*There are 9.2% missing BMI entries (n=1010). \*\* There are 3.6% missing Parity entries (n=394).

### Performance of polygenic risk score in predicting GDM

The performance of the polygenic risk score in predicting GDM is shown in Figure 2. The PRS z-score was higher in women with GDM compared with controls (mean z-score GDM 0.34 ± 0.95 vs Controls -0.14 ± 0.99; p < 2.2 × 10¹). British Bangladeshi women (mean z-score GDM 0.19 ± 0.89 vs Controls 0.52 ±0.89; p = 2.2×10^−16^) had higher z-scores compared to Pakistani women (mean z-score GDM -0.24 ±0.89 vs Controls -0.63 ±0.92; p = 2.2×10^−16^). A histogram depicting the distribution of the PRS across the two groups can be found in (Supplementary Figure 2).

**Figure 2.**
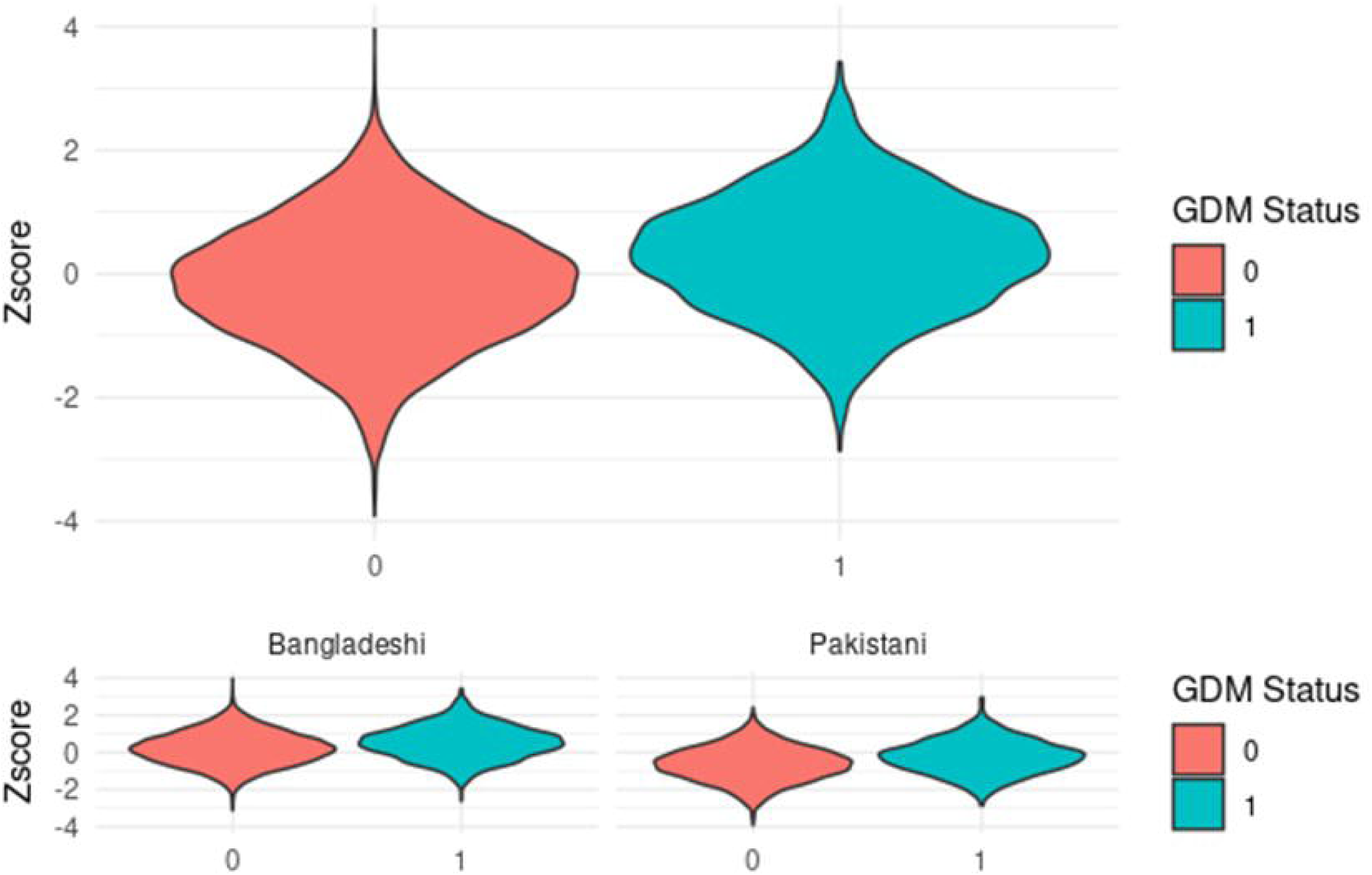
Polygenic risk score distribution amongst women with and without GDM. GDM Status: 0 = women who did not have GDM; 1 = women who had GDM. Abbreviations: GDM – gestational diabetes.

### Association between PRS and risk of developing GDM

To further investigate the relationship between the PRS and GDM risk, we divided our participants into deciles based on their PRS values. As demonstrated in figure 3, the likelihood of diabetes increased incrementally with increasing PRS deciles. The odds of having gestational diabetes in the 10^th^ decile compared to the 1^st^ decile were 5.66 (95% CI = [4.59, 7.01], p = 3.62 x 10^-58^). These results were replicated in the subcohorts with British Bangladeshi women in the highest risk decile being 3.8 (95% CI = [3.01, 4.82], p = 1.14 x 10^-34^) and Pakistani women in the highest risk decile 5.03 (95% CI = [3.42, 7.54], p = 9.14 x 10^-15^) more likely to develop GDM, compared to those in the lowest risk decile (Supplementary Figure 3).

**Figure 3.**
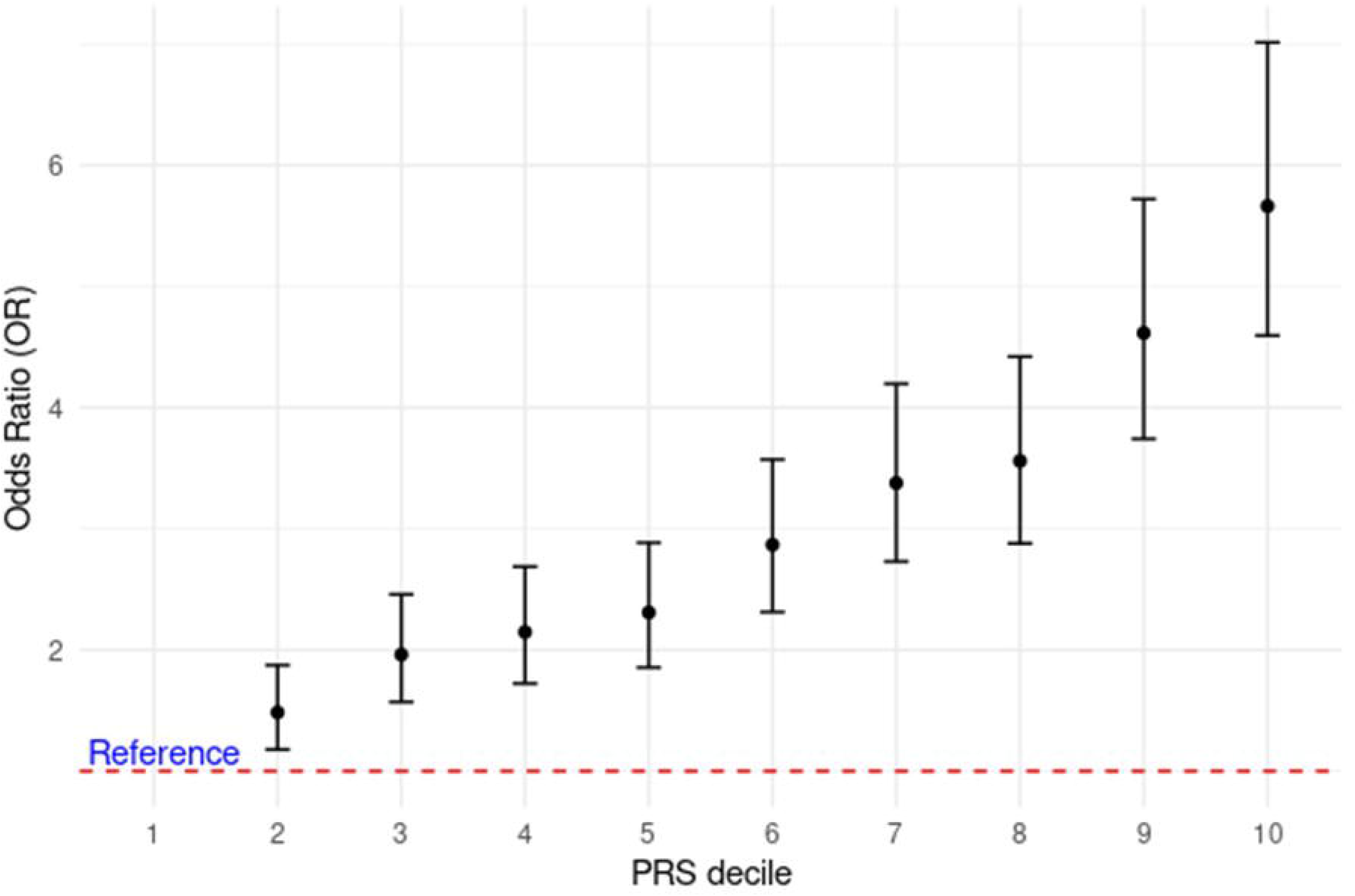
Risk of developing gestational diabetes by polygenic risk score decile. Abbreviations: PRS – polygenic risk score.

### Comparison of null (population risk factors only), genetic and combined models in predicting gestational diabetes using AUC

The receiver operating curve (ROC; Figure 4) demonstrates that combining population and genetic risk improves comparison of gestational diabetes when compared to the null model using population risk factors only. There is an improvement in the C-statistic from 0.66 (95% CI = [0.65, 0.67]) to 0.70 (95% CI = [0.68, 0.71]); this improvement is statistically significant with a p-value of < 2.2 × 10¹. These results were also replicated in stratified subcohorts of British Bangladeshi (Full model AUC = 0.68, 95% CI = [0.66–0.69] vs null model AUC = 0.64, 95% CI = [0.62-0.65]), p = 2.2 × 10¹ and Pakistani women (Full model AUC = 0.69, 95% CI = [0.67–0.71] vs null model AUC = 0.65, 95% CI = [0.62-0.67], p = 0.14 x 10 x ^-8^).

**Figure 4.**
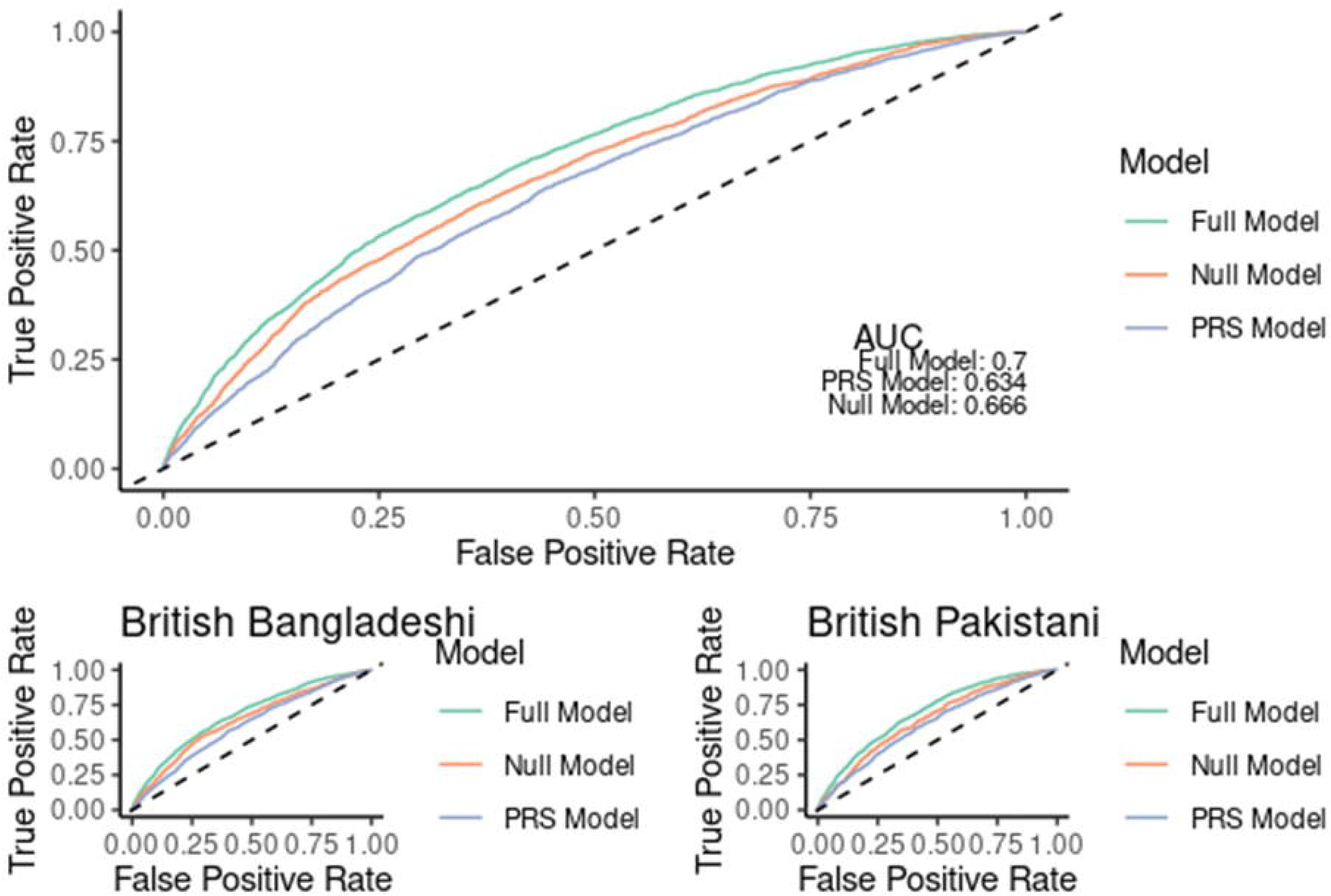
ROC curves demonstrating improvement in prediction of GDM with utilization of genetic information. Comparing three models: the Null model, which includes only traditional population risk factors (age, BMI, parity, and ethnicity); the PRS model, which includes only the polygenic risk score; and the Full model, which combines both the population risk factors and the polygenic risk score. Abbreviations: AUC – area under the curve; PRS – polygenic risk score; ROC – receiver operating characteristic.

### Performance of polygenic risk score in predicting transfer from GDM to type 2 diabetes

The performance of the polygenic risk score in predicting development of type 2 diabetes following GDM is shown in Figure 5. The PRS z-score was higher in women with GDM who progressed to type 2 diabetes (T2DM) compared with those that did not go on to develop T2DM (mean z-score GDM to T2DM 0.27 ± 0.97 vs no progression to T2DM -0.06 ± 0.99; p < 6.704 × 10^14^). British Bangladeshi women (mean z-score GDM to T2DM 0.27 ± 0.97 vs no progression to T2DM - 0.06 ± 0.99; p = 4.169 x 10^-11^) had lower z-scores compared to Pakistani women (mean z-score GDM toT2DM 0.28 ±0.98 vs no progression to T2DM -0.05 ±0.99; p = 0.00026). Individuals with a previous history of gestational diabetes in the highest decile were 3.44 times more likely to develop type 2 diabetes.

**Figure 5.**
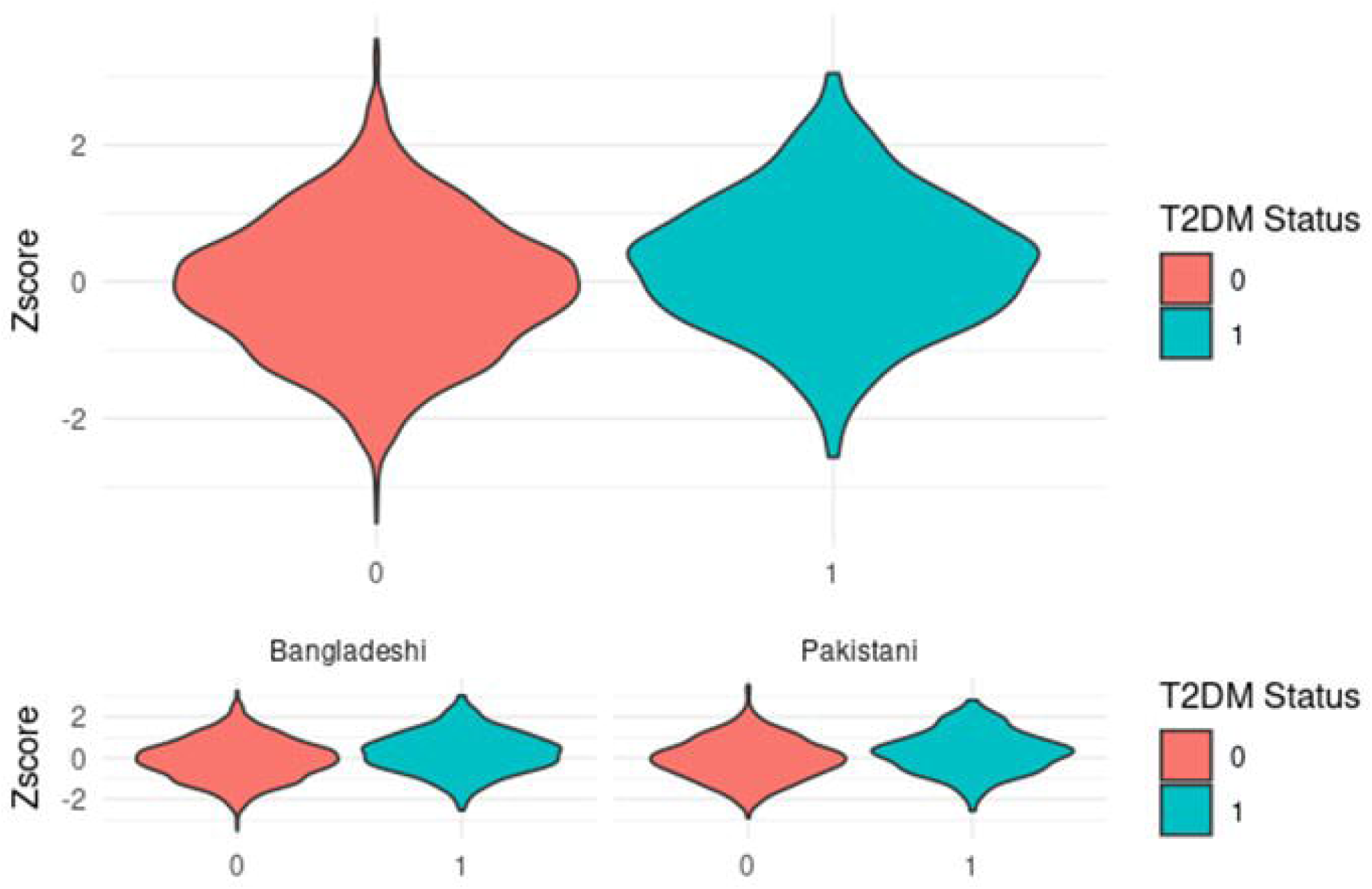
Polygenic risk score distribution amongst women with GDM who progressed to type 2 diabetes versus those who did not. T2DM Status: 0 = women who did progress to type 2 diabetes; 1 = women who progressed to type 2 diabetes. Abbreviations: T2DM –type 2 diabetes.

Over 10 years, there was no significant difference between days to progression to type 2 diabetes between the first and highest decile (median days highest decile 1213 [IQR: 646 - 2073] compared to lowest decile 1562 [IQR: 828 - 2863], p = 0.141; Supplementary Figure 4). There was no evidence of younger age at the time of type 2 diabetes onset between the highest and lowest decile (Supplementary Figure 5). Table 2 demonstrates the percentage prediction of risk of conversion from gestational diabetes to type 2 diabetes.

**Table 2.** Percentage prediction of risk of conversion from gestational diabetes to type 2 diabetes, based on combined model incorporating polygenic risk score and population risk factors (age, parity, BMI).

| Prediction of risk | Conversion to Type 2 diabetes |
| --- | --- |
| Lowest decile of predicted risk | 11% |
| All GDM cases | 19% |
| Highest decile of predicted risk | 30% |

Abbreviations: GDM – gestational diabetes.

### Comparison of null (population risk factors only), genetic and combined models in predicting development of type 2 diabetes following gestational diabetes using AUC

The receiver operating curve (ROC; Figure 6) demonstrates an improvement in performance by addition of genetic risk information to population factors alone. Specifically, the c-statistic was improved from 0.62 (null model 95% CI = [0.60, 0.65]) to 0.67 (full model 95% CI = [0.65, 0.69]), with a p-value of = 4.583 x 10^-6^. These results were also replicated across both stratified sub-cohorts of British Bangladeshi (Full model AUC = 0.66, 95% CI = [0.63–0.69] vs null model AUC = 0.60, 95% CI = [0.58-0.63]) and Pakistani women (Full model AUC = 0.75, 95% CI = [0.71–0.80] vs null model AUC = 0.71, 95% CI = [0.67-0.76], p = 0.0088).

**Figure 6.**
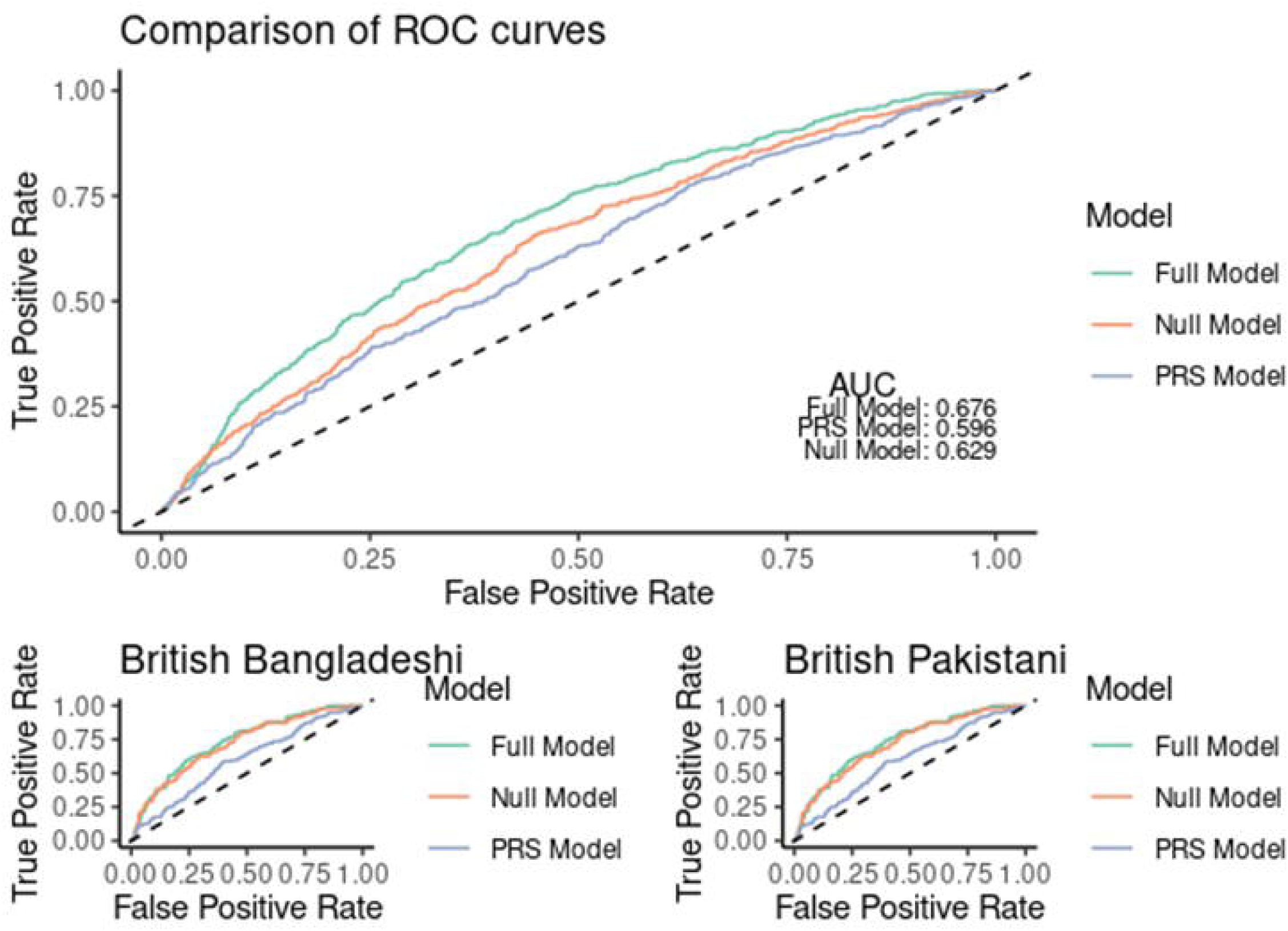
ROC curves demonstrating improvement in prediction of type 2 diabetes with utilization of genetic information. Comparing three models: the Null model, which includes only traditional population risk factors (age, BMI, parity, and ethnicity); the PRS model, which includes only the polygenic risk score; and the Full model, which combines both the population risk factors and the polygenic risk score. Abbreviations: AUC – area under the curve; PRS – polygenic risk score; ROC – receiver operating characteristic.

## Comment

### Principal Findings

In this study of 10,931 women of South Asian ethnic origin, the addition of genetic information to epidemiological risk factors offered some improvement to the prediction of both primary gestational diabetes, and the subsequent development of type 2 diabetes following GDM. By combining epidemiological and genetic risk factors, it is possible to identify women at a substantially higher risk than background (30%) of developing type 2 diabetes following GDM, offering a possible cohort for therapeutic intervention.

### Results in the Context of What is Known

It has previously been demonstrated that incorporating genetic information can improve the prediction of type 2 diabetes after GDM [5]. However, the potential contribution of genetics has not been fully explained. In our cohort, we were able to stratify women based on their genetic risk, with women in the highest risk group at 1.6 times the risk of the group overall (30% compared to 19%) and women in the lowest risk group at half the risk (10% compared to 19%).

The underlying etiology of type 2 diabetes is multifactorial, including genetic, epigenetic, environmental, and nutritional factors. Both type 2 and gestational diabetes are diagnoses subject to changing definition, with the threshold of diagnosis for each coming down in recent years,[17] based on heterogenous evidence with different endpoints in mind and on different diagnostic strategies. The two pathologies are not exactly analogous and there remain outstanding questions about both underlying genotypes and phenotypes of both. This study indicates that, in this South Asian cohort, underlying genetic susceptibility has the potential to clarify this understanding.

### Strengths and Limitations

This study’s main strength lies in the data source: this is a large population-based study of women at high underlying risk of gestational and type 2 diabetes with linked health and genomic data to improve the prediction of diabetes.

This study is limited by the relative homogeneity of the cohort, potentially limiting translation into wider populations; further analysis and external validation will be required to understand the implications for women from other ethnic groups. This study is further limited by data availability, including the need to impute data missing in electronic health records. It is possible that additional clinical information may offer further improvement of prediction.

### Clinical and Research Implications

This study demonstrates the feasibility of using genetic information to identify women at higher risk of type 2 diabetes after GDM. Identifying women at very high risk for ongoing diabetes after gestational diabetes would provide a cohort for investigation of ongoing intervention in the immediate postnatal period and prioritization for more intensive postnatal monitoring, to improve outcomes both for future pregnancies and lifelong. This will require additional development and external validation.

This study also demonstrates the small potential use of genetic information to improve prediction of gestational diabetes in this high-risk cohort. In practice, women from South Asian ethnic groups are universally recommended to have screening for gestational diabetes at 24-28 weeks of gestation in high-income settings [13,18] and, given the severity of a missed diagnosis of gestational diabetes, this is unlikely to be altered unless genetic information could be used to rule-out gestational diabetes.

This suggests that genetic studies in gestational diabetes should not be focused on identification in this cohort but rather on understanding disease trajectory, treatment strategies, and future risk, including recurrence in women planning subsequent pregnancy.

## Conclusions

This study, in a large cohort of women from British Bangladeshi and Pakistani backgrounds, demonstrates the feasibility and potential utility of genetic information combined with epidemiological risk factors to identify women at high risk of type 2 diabetes after gestational diabetes. Further research should focus on the development of risk prediction tools and evaluation of opportunities for intervention in higher-risk cohorts.

## Data Availability

All data produced in the present study are available upon reasonable request to the authors.

## Guarantor of the article

Dr Julia Zöllner PhD MRCOG

## Patient consent

ELGH (East London Genes and Health) operates under ethical approval, 14/LO/1240, from London South East NRES Committee of the Health Research Authority, dated 16 September 2014. This protocol was approved by ELGH on 5^th^ July 2023, reference S00099. No further patient consent is required to utilize this anonymized data set.

## Specific author contributions

JZ and SF conceived the idea of the study. JJ and JZ applied for data access. JZ accessed, extracted, and verified the data. JZ and JJ analyzed and interpreted the data. JJ and JZ wrote the first draft of the manuscript, and all other authors contributed to subsequent drafts. BO, SH, SF, SI, MZ, RM, and DvH contributed to data interpretation. All authors gave final approval of the final draft to be published and have contributed to the manuscript.

## Condensation page

### Tweetable statement

Women with gestational diabetes, especially of South Asian descent, are at high risk for type 2 diabetes. Combining genetic and demographic factors can improve risk prediction and inform targeted interventions.

### Short Tile

Genetic risk scores improve prediction for gestational and type 2 diabetes in British Pakistani and Bangladeshi women.

## Acknowledgements

We thank Social Action for Health, Centre of The Cell, members of our Community Advisory Group, and staff who have recruited and collected data from volunteers. We thank the NIHR National Biosample Centre (UK Biocentre), the Social Genetic & Developmental Psychiatry Centre (King’s College London), Wellcome Sanger Institute, and Broad Institute for sample processing, genotyping, sequencing and variant annotation.

This work uses data provided by patients and collected by the NHS as part of their care and support. This research utilised Queen Mary University of London’s Apocrita HPC facility, supported by QMUL Research-IT, http://doi.org/10.5281/zenodo.438045

We thank: Barts Health NHS Trust, NHS Clinical Commissioning Groups (City and Hackney, Waltham Forest, Tower Hamlets, Newham, Redbridge, Havering, Barking and Dagenham), East London NHS Foundation Trust, Bradford Teaching Hospitals NHS Foundation Trust, Public Health England (especially David Wyllie), Discovery Data Service/Endeavour Health Charitable Trust (especially David Stables), Voror Health Technologies Ltd (especially Sophie Don), NHS England (for what was NHS Digital) - for GDPR-compliant data sharing backed by individual written informed consent.

Most of all we thank all of the volunteers participating in Genes & Health.

*Current <u>Genes & Health Research Team</u> (in alphabetical order by surname): Shaheen Akhtar^4^, Mohammad Anwar^5^, Omar Asgar^6^, Samina Ashraf^7^, Saeed Bidi^8^, Gerome Breen^9^, James Broster^8^, Raymond Chung^9^, David Collier^8^, Charles J Curtis^9^, Shabana Chaudhary^8^, Grainne Colligan^5^, Panos Deloukas^8^, Ceri Durham^5^, Faiza Durrani^8^, Fabiola Eto^8^, Sarah Finer^3^, Joseph Gafton^8^, Ana Angel^8^, Chris Griffiths^8^, Joanne Henry^8^, Teng Heng^4^, Sam Hodgson^3^, Qin Qin Huan^4^, Matt Hurles^4^, Karen A Hunt^8^, Shapna Hussain^8^, Kamrul Islam^8^, Vivek Iyer^4^, Benjamin M Jacobs^8^, Georgios Kalantzis^4^, Ahsan Khan^8^, Claudia Langenberg^8^, Cath Lavery^8^, Sang Hyuck Lee^9^, Daniel MacArthur^11^, Eamonn Maher^10^, Sidra Malik^8^, Daniel Malawsky^4^, Hilary Martin^4^, Dan Mason^12^, Rohini Mathur^8^, Mohammed Bodrul Mazid^8^, John McDermott^13^, Caroline Morton^8^, Bill Newman^13^, Vladimir Ovchinnikov^8^, Elizabeth Owor^8^, Iaroslav Popov^8^, Asma Qureshi^8^, Mehru Raza^8^, Jessry Russell^8^, Nishat Safa^8^, Miriam Samuel^8^, Moneeza Siddiqui^3^, Michael Simpson^9^, John Solly^8^, Marie Spreckley^8^, Daniel Stow^8^, Michael Taylor^8^, Richard C Trembath^9^, Karen Tricker^8^, David A van Heel^2^, Klaudia Walter^4^, Caroline Winckley^14^, Suzanne Wood^8^, John Wright^15^, Sabina Yasmin^8^, Ishevanhu Zengeya^8^, Julia Zöllner^1,3^.

^4^Wellcome Sanger Institute, London, UK. ^5^Social Action for Health, London, UK. ^6^Manchester University NHS Trust, Manchester, UK. ^7^Bradford Teaching Hospitals, Bradford, UK. ^8^Queen Mary University of London, London, UK. ^9^King’s College London, London, UK. ^10^University of Cambridge, Cambridge UK. ^11^Garvan Institute of Medical Research, Darlinghurst, NSW, Australia. ^12^Born in Bradford, Bradford, UK. ^13^University of Manchester, Manchester, UK. ^14^NIHR Clinical Research Clinical Trials, Manchester, UK. ^15^Bradford Institute for Health Research, Bradford, UK.

## Supplementary Documentation

**Supplementary Methods. Description of quality control, imputation of genotype data, as well as filtering and principal component analysis.**

**Supplementary Table 1. Table of included SNOMED or ICD-10 codes.**

**Supplementary Figure 1. Comparison of different PRS performance with predicting GDM (beta >0.3).** Abbreviations: GDM – gestational diabetes; PRS – polygenic risk score.

**Supplementary Figure 2. Distribution of PRS (z-score) by GDM status.**

Abbreviations: GDM – gestational diabetes; PRS – polygenic risk score.

**Supplementary Figure 3. Risk of developing GDM by PRS decile divided by different genetic ancestry.** Abbreviations: GDM – gestational diabetes; PRS – polygenic risk score.

**Supplementary Figure 4. Progression (days) to type 2 diabetes between top and bottom deciles.**

**Supplementary Figure 5. Age distribution among bottom and top decile of PRS.** Abbreviations: PRS – polygenic risk score.

