## Supplementary figures and images for "Understanding the potential contribution of polygenic risk scores to the prediction of gestational and type 2 diabetes in women from British Pakistani and Bangladeshi groups: a cohort study in Genes and Health"

### Supplemental Figure 1

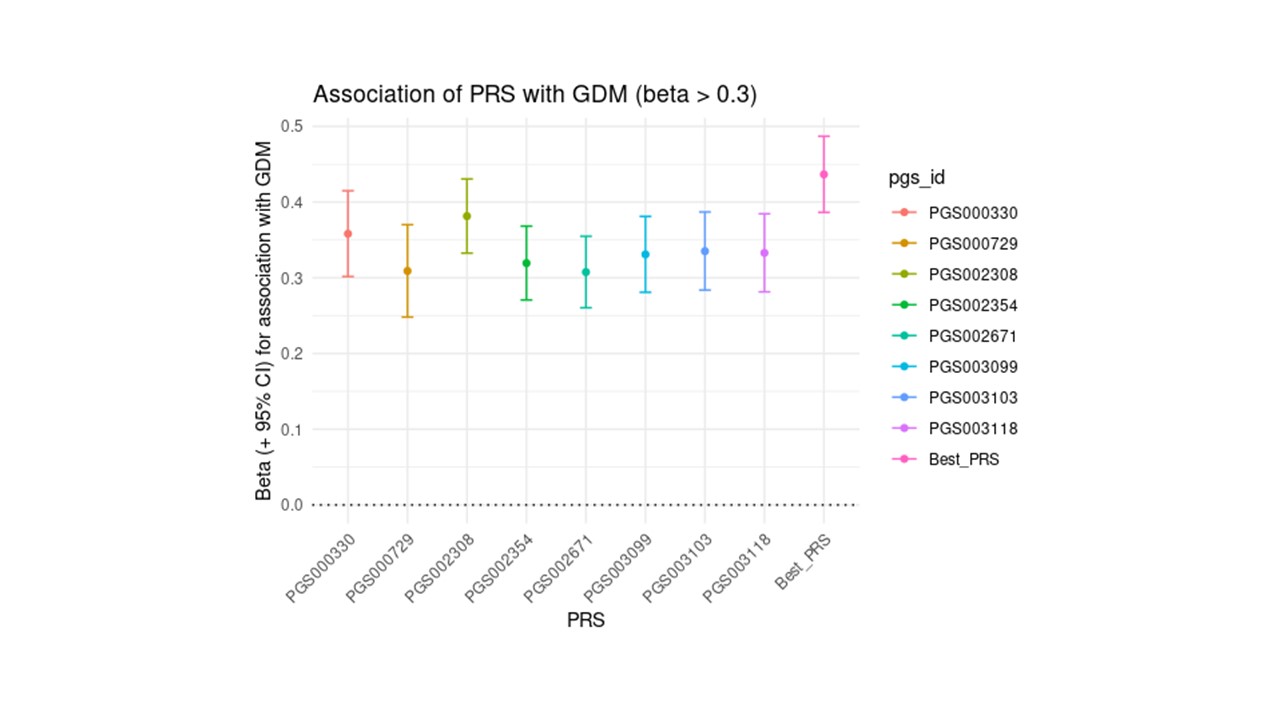

### Supplemental Figure 2

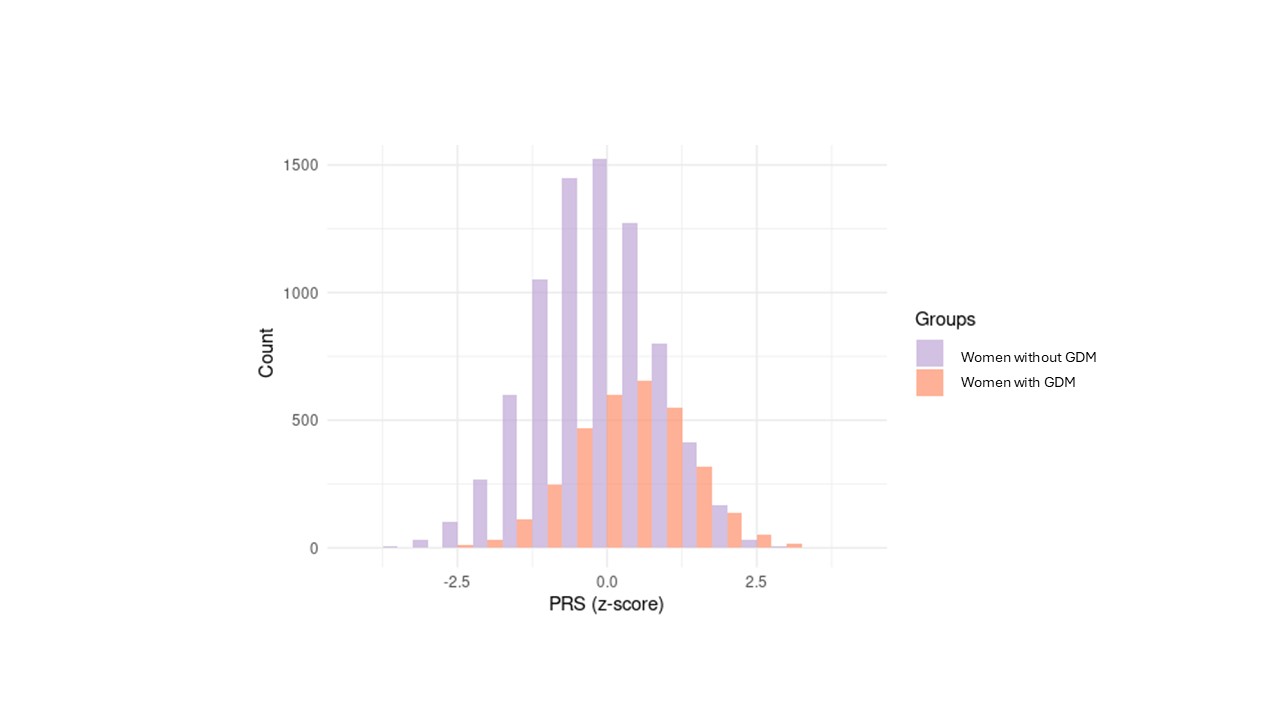

### Supplemental Figure 3

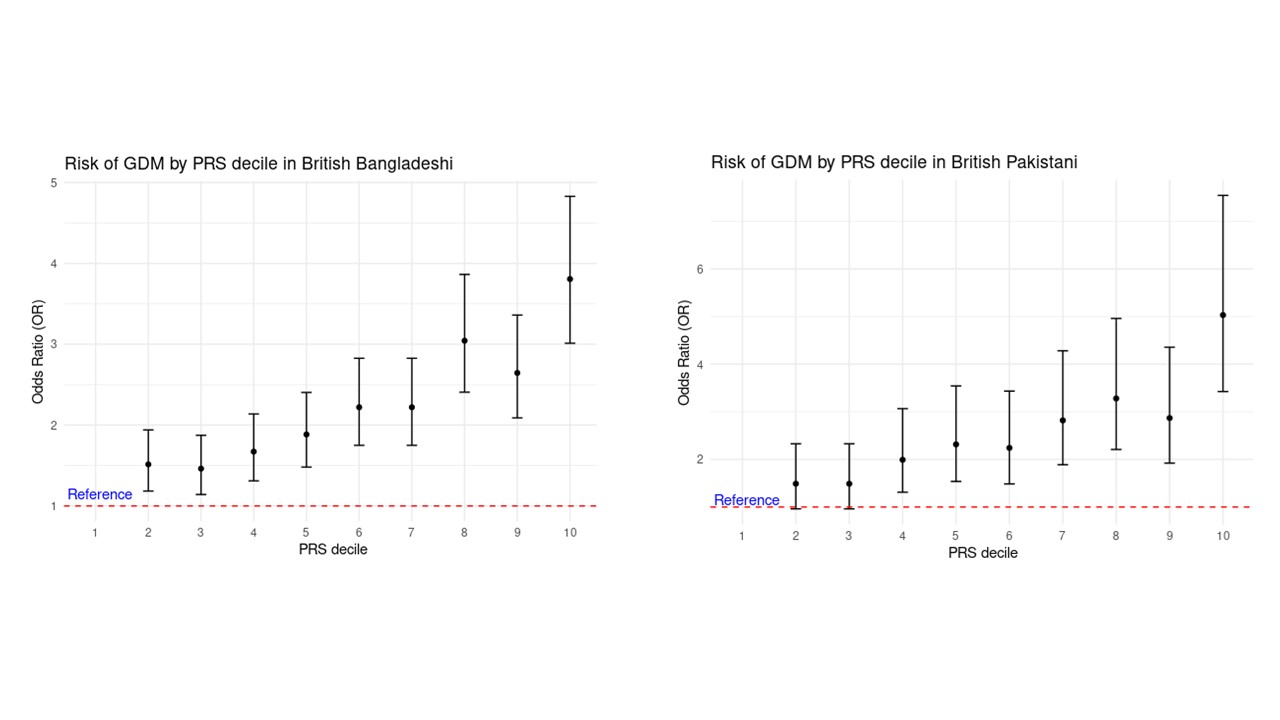

### Supplemental Figure 4

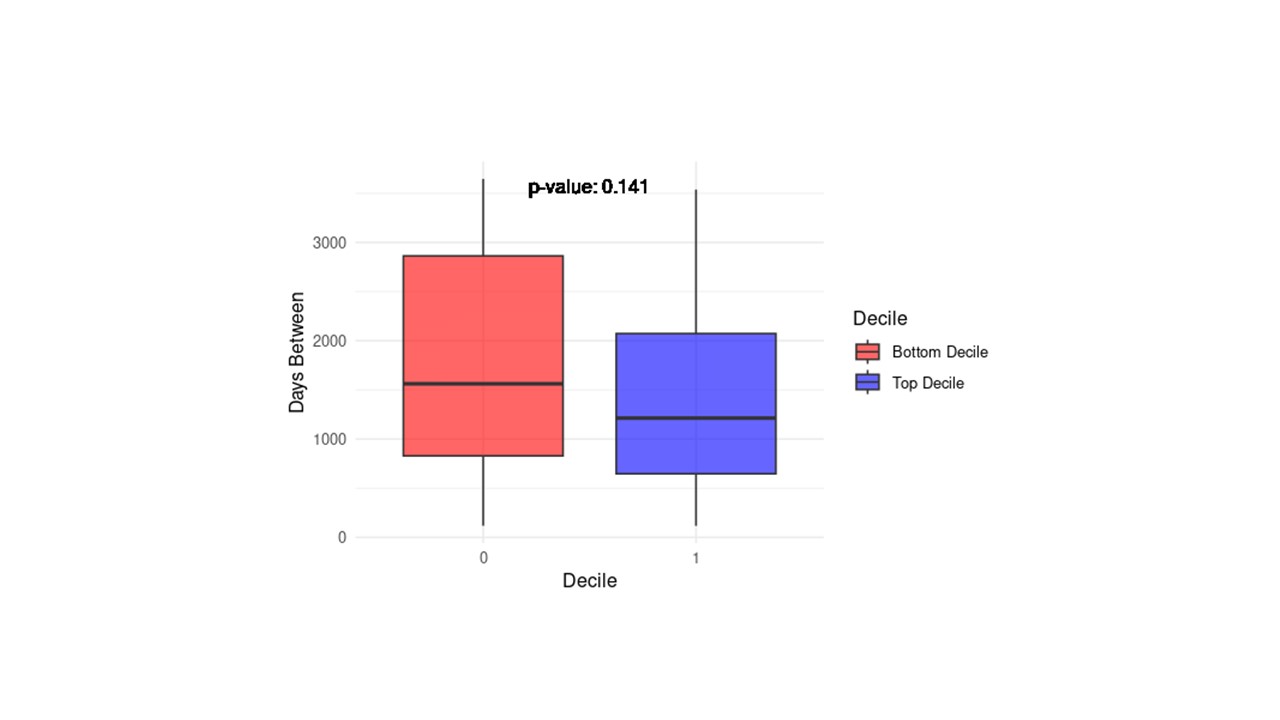

### Supplemental Figure 5

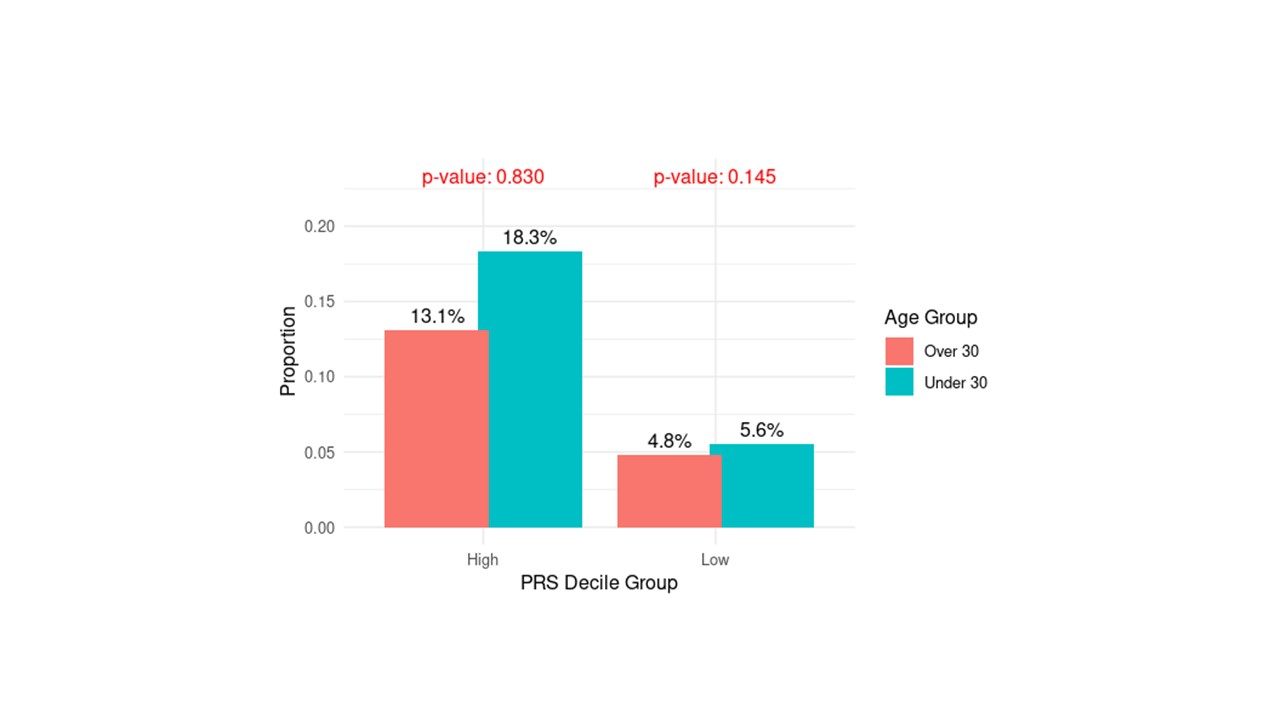
