## Supplemental Methods for "Understanding the potential contribution of polygenic risk scores to the prediction of gestational and type 2 diabetes in women from British Pakistani and Bangladeshi groups: a cohort study in Genes and Health"

**Supplementary Methods**

**Genotyping**

DNA was extracted from saliva samples collected using Oragene saliva sampling kits. Individuals were genotyped using the Illumina Global Screening Array version 3 (GSA v3) chip supplemented with additional multi-disease content. Initial genotype calling and quality control were performed with Illumina GenomeStudio version 2.0. Specifically, automated clustering via the GenTrain algorithm was applied to 1,970 selected high-quality samples at a subset of high-confidence variants—autosomal variants in Hardy–Weinberg equilibrium with GenTrain scores exceeding 0.7. Multiple rounds of manual and automated reclustering were then conducted to identify and exclude low-quality variants and samples. This process resulted in a dataset achieving over 99% call rate across 637,829 single-nucleotide polymorphisms (SNPs). The established cluster file was subsequently used to genotype the remaining approximately 50,000 samples. Samples were excluded if their call rates per sex were lower than those in the initial batch (<99.2% for females, <99.5% for males). From the initial 54,206 genotyped samples, individuals were removed due to missing NHS numbers, discrepancies between reported gender and genetic sex, or implausible genetic duplicates (i.e., duplicates not explained by twin status). This filtering yielded a dataset of 51,176 individuals genotyped at 608,329 autosomal SNPs with a genotyping rate exceeding 99.9%.

Following the exclusion of rare variants (minor allele frequency [MAF] < 0.0001), palindromic variants, and insertions/deletions (indels), genotypes were imputed to the TOPMed r3 multi-ancestry imputation panel aligned to genome build hg38 using the TOPMed Imputation Server^1^. Post-imputation, we conducted single-nucleotide polymorphism (SNP) quality control by filtering for common (MAF > 0.01) biallelic autosomal variants with less than 10% missingness, imputation quality scores (INFO) greater than 0.7, and no significant deviation from Hardy–Weinberg equilibrium (P > 1 × 10^-15^). Variants with duplicate genomic positions were removed. Imputed genotype dosages falling outside the ranges of 0–0.1, 0.9–1.1, or 1.9–2.0 were set to missing to ensure data integrity. Individuals with more than 10% missing genotypes were excluded from further analysis. Genetic duplicate samples were identified using KING^2^; specifically, 10 pairs of probable identical twins were detected, and one individual from each pair was removed to avoid redundancy. Principal component analysis was utilized to identify and exclude fewer than 10 ancestral outliers who did not cluster with South Asian reference samples from the Human Genome Diversity Project and the 1000 Genomes Project^3^. Through a clustering-based procedure to estimate categorical ancestry groupings (Bangladeshi or Pakistani), an additional 62 participants were excluded due to ambiguous ancestry assignments. We further excluded participants who had missing covariate information (age and sex) or were not linked to electronic healthcare records.
